# Prompt Engineering Strategies Improve the Diagnostic Accuracy of GPT-4 Turbo in Neuroradiology Cases

**DOI:** 10.1101/2024.04.29.24306583

**Authors:** Akihiko Wada, Toshiaki Akashi, George Shih, Akifumi Hagiwara, Mitsuo Nishizawa, Yayoi Hayakawa, Junko Kikuta, Keigo Shimoji, Katsuhiro Sano, Koji Kamagata, Atsushi Nakanishi, Shigeki Aoki

## Abstract

**Background:** Large language models (LLMs) like GPT-4 demonstrate promising capabilities in medical image analysis, but their practical utility is hindered by substantial misdiagnosis rates ranging from 30-50%.

**Purpose:** To improve the diagnostic accuracy of GPT-4 Turbo in neuroradiology cases using prompt engineering strategies, thereby reducing misdiagnosis rates.

**Materials and Methods:** We employed 751 publicly available neuroradiology cases from the American Journal of Neuroradiology Case of the Week Archives. Prompt instructions guided GPT-4 Turbo to analyze clinical and imaging data, generating a list of five candidate diagnoses with confidence levels. Strategies included role adoption as an imaging expert, step-by-step reasoning, and confidence assessment.

**Results:** Without any adjustments, the baseline accuracy of GPT-4 Turbo was 55.1% to correctly identify the top diagnosis, with a misdiagnosis rate of 29.4%. Considering the five candidates’ improved applicability, it is 70.6%. Applying a 90% confidence threshold increased the accuracy of the top diagnosis to 72.9% and the applicability of the five candidates to 85.9%, while reducing misdiagnoses to 14.1%, but limited the analysis to half of cases.

**Conclusion:** Prompt engineering strategies with confidence level thresholds demonstrated the potential to reduce misdiagnosis rates in neuroradiology cases analyzed by GPT-4 Turbo. This research paves the way for enhancing the feasibility of AI-assisted diagnostic imaging, where AI suggestions can contribute to human decision-making processes. However, the study lacks analysis of real-world clinical data. This highlights the need for further investigation in various specialties and medical modalities to optimize thresholds that balance diagnostic accuracy and practical utility.

## Introduction

Large language models (LLMs) have shown considerable promise in the processing of textual information, achieving performance levels that approximate human expertise (1–8). These models have been applied in various fields, including medicine, where they offer the potential to assist in the interpretation of complex medical data. In medical imaging diagnostics, LLMs, such as GPT-3.5 and GPT-4, have been explored for their ability to analyze radiological texts. Current reports suggest that these models achieve approximately 50-69% accuracy in identifying correct diagnoses from imaging findings (9–13). However, the 30-50% misdiagnosis rate is relatively high, which may be insufficient for assisting healthcare professionals in making accurate clinical decisions. Physicians generally seek diagnostic tools that minimize misdiagnosis risks to ensure patient safety and effective treatment. This potential and limitations of LLM in medical applications and the need to improve accuracy and reduce misdiagnosis rates in clinical settings (14).

Prompt engineering, a method of giving precise instructions to LLMs, has shown effectiveness in eliciting desired responses and is reported to improve LLMs’ performance in various applications (15). In this study, we explore how prompt engineering can improve the diagnostic accuracy of LLM in medical imaging to reduce misdiagnosis rates. In this study, we aim to utilize the latest LLM, GPT-4 Turbo, along with prompt engineering techniques to improve diagnostic abilities in medical imaging, particularly focusing on reducing misdiagnosis rates. By adopting these advanced technologies, we seek to improve the precision of diagnostic suggestions, using the capabilities of GPT-4 Turbo to address the challenges of accurately interpreting medical images.

## Materials and Methods

This study was carried out using the checklist for artificial intelligence in medical imaging. It was exempted from institutional review board oversight because it used publicly available data.

### Data Collection

Our methodology examined 751 publicly available neuroradiological cases from 2012 to 2023 from the American Journal of Neuroradiology (AJNR) Case of the Week Archives (https://www.ajnr.org/cow/by/diagnosis) (16). The AJNR Case of the Week site separates clinical information, images, and diagnoses/explanations into separate tabs. GPT-4 Turbo accessed the indicated URL to retrieve clinical information and image findings as textual information without knowing the diagnosis name.

### AI Model and Platform

We leveraged GPT-4 Turbo, specifically the “gpt-4-1106-preview” model, an advanced version of LLM developed by OpenAI (17). This version, notable for its enhanced capabilities and a vast 128k context window, allows for comprehensive analysis within a single prompt. We use the MD.ai Reporting/Chat application (https://chat.md.ai?) for direct URL access to extract relevant clinical and imaging information, omitting diagnoses to test the diagnostic accuracy of GPT-4 Turbo.

### Prompt Instruction

In this study, we used three strategy prompt designs to improve the precision of GPT -4 turbo diagnostic suggestions in neuroradiology cases. These strategies include role adoption, step-by-step thinking, and confidence level assessment (Figure 1). We introduced these strategies based on the knowledge from recent engineering guides and reports (15,18).

**Figure 1:**
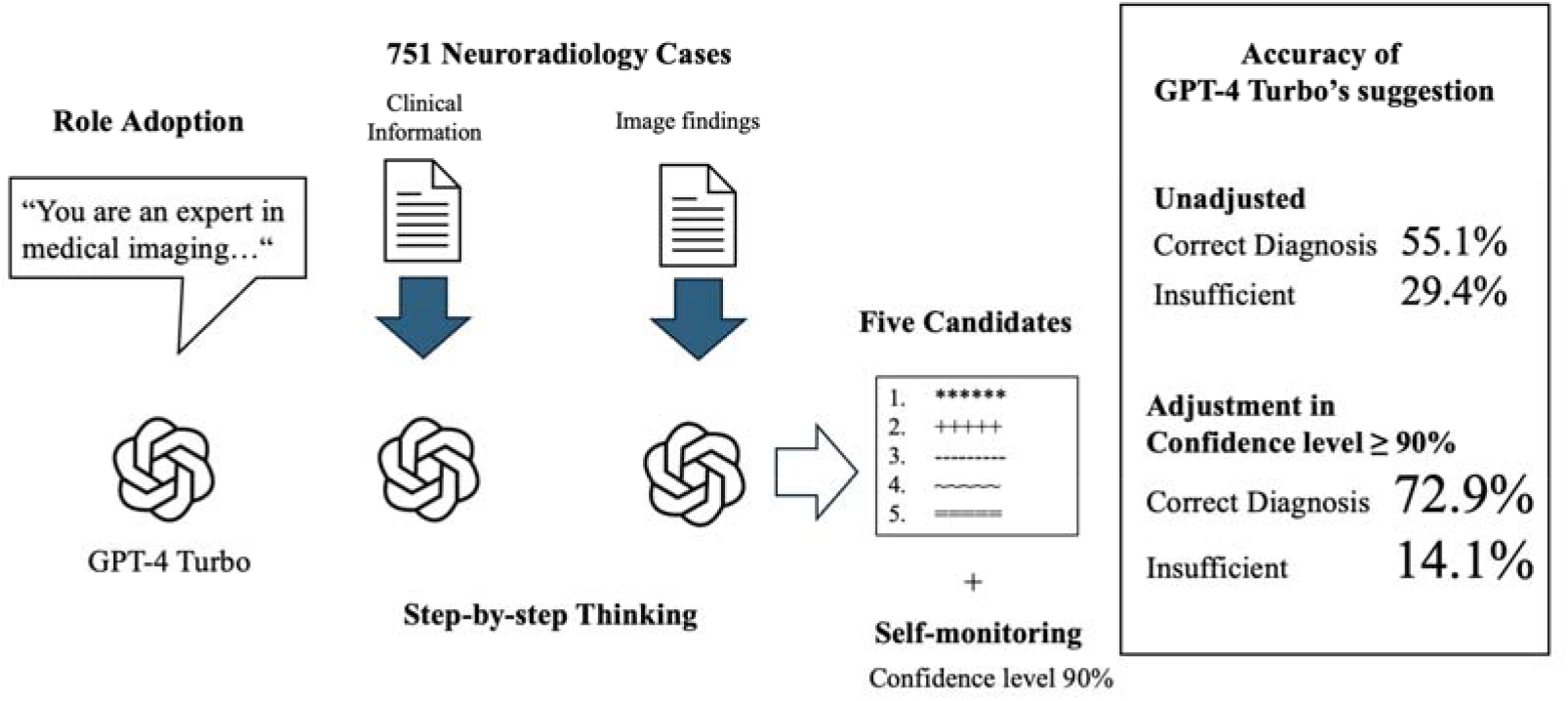
Methodology for prompt engineering to enhance GPT-4 Turbo’s utility and reduce misdiagnoses in interpreting 751 neuroradiology cases from the American Journal of Neuroradiology Case of the Week Archives. A role-adoption prompt made GPT-4 Turbo an expert radiologist. GPT-4 Turbo extracted textual clinical information and imaging findings and then used step-by-step thinking to generate a list of 5 diagnostic candidates with confidence levels. The unadjusted performance showed a 55.1% correct diagnosis and 29.4% insufficient suggestions. Adjusting for a confidence level threshold of 90% improved the correct diagnosis to 72.9% but with 14.1% insufficient suggestions on the reduced subset meeting that threshold.

### Role Adoption

Adopting roles involves directing the LLM to act as an expert in the diagnosis of medical imaging. This strategy aims to shift the LLM’s process from data processing to decision-making with domain knowledge. By acting as an expert, the LLM can prioritize relevant information for disease diagnosis from medical images such as CT, MRI, and X-rays. This approach is based on the assumption that by narrowing the LLM’s focus to a specific domain, the quality and relevance of its diagnostic suggestions will improve. Role-specific data extraction will lead to higher response precision as the LLM tailors its output to reflect the experience expected of a medical imaging diagnostician.

### Step-by-Step Thinking

Step-by-step thinking requires the LLM to take a systematic approach to the diagnostic task, mirroring the logical progression a human expert would take in analyzing clinical information and imaging findings. Our prompt instructs the LLM to suggest an initial differential diagnosis from clinical information alone, then receive imaging findings, and modify the differential diagnosis. This approach encourages the LLM to gradually integrate and refine candidate diagnoses with input data. Such a structured approach could make LLM diagnostic reasoning more transparent and more accessible to interpret, potentially increasing the accuracy and reliability of the suggestions provided.

### Confidence assessment

Instructions that ask LLMs to analyze their responses and evaluate their confidence level are called “self-monitoring.”(18) LLMs self-assess their confidence based on task difficulty, match with training data, and consistency of their responses. Users can obtain quantitative information about the LLM’s list of candidate diagnoses.

### The specific text of the instructions for GPT-4 Turbo

The following prompts enabled GPT-4 Turbo to extract textual data of clinical information and imaging findings from the AJNR case of the week site and generate a list of diagnostic candidates and their confidence levels.

**# Role**

You are an expert in medical imaging diagnosis with extensive experience interpreting various medical images, including CT, MRI, and X-rays. Your expertise includes identifying pathologies, understanding radiology clinical report contexts, and correlating to imaging findings with potential diagnoses proofread.

**# Request**

Along with the following Regulation prompt, present a refined list of five differential diagnoses, including the most probable diagnosis and four alternatives.

Each diagnosis should have a corresponding confidence level based on your comprehensive analysis.

**# Regulation**

Using the clinical information provided: {# URL of clinical information}, list five initial differential diagnoses. Then, review the imaging findings: {# URL of image findings}, and update your diagnoses accordingly. Reflect on how the new data alters your assessment. For each diagnosis in your updated list, assign a confidence level between 0% and 100%, considering the task’s complexity and the extent to which clinical and imaging data support each diagnosis.

### Evaluation of GPT-4 Turbo’s Diagnostic Accuracy

Two board-certified neuroradiologists with 15 and 28 years of clinical experience (T.A. and A.W) evaluated the diagnostic suggestions from GPT-4 Turbo. We evaluated the accuracy of GPT-4 Turbo’s responses using a three-tier scale: “Excellent” for instances where the top diagnostic suggestion matched the correct diagnosis, “Good” when the correct diagnosis was among the suggested candidates, and “Insufficient” if the correct diagnosis was not listed among the suggested candidates.

## Results

Table 1 presents the breakdown of the diagnostic performance of GPT-4 Turbo in 751 cases of neuroradiology. The ‘Excellent’ category, where the top predicted diagnosis matched the ground truth, was achieved in 55.1% of the cases. The ‘Good’ category, where the correct diagnosis was included among the top five predictions, covered an additional 15.5% of cases, totaling 70.6%. However, in 29.4% of cases categorized as ‘Insufficient,’ the correct diagnosis was not present within GPT-4 Turbo’s predictions.

**Table 1.** Breakdown of diagnostic tasks of neuroradiological imaging. The ‘Excellent’ category represents cases where the top diagnostic prediction matches the correct diagnosis. ‘Good’ indicates cases where the correct diagnosis was included within the top five diagnostic predictions. ‘Insufficient’ marks cases where the correct diagnosis was not included in the diagnostic predictions.

| **Disease Category** | **Excellent** | **Good** | **Insufficient** | **Proportion of Total Cases** |
| --- | --- | --- | --- | --- |
| Tumor | 0.442 | 0.191 | 0.367 | 0.29 (215) |
| Demyelinating Disease | 0.727 | 0.000 | 0.273 | 0.01 (11) |
| Infections and Inflammatory Disease | 0.535 | 0.153 | 0.313 | 0.19 (144) |
| Vascular Disease | 0.688 | 0.150 | 0.163 | 0.11 (80) |
| Genetic/Degenerative Disease | 0.515 | 0.134 | 0.351 | 0.13 (97) |
| Trauma | 0.667 | 0.267 | 0.067 | 0.02 (15) |
| Metabolic Disease | 0.735 | 0.088 | 0.176 | 0.09 (68) |
| Malformation | 0.563 | 0.229 | 0.208 | 0.06 (48) |
| Neurological Disease | 0.571 | 0.000 | 0.429 | 0.01 (7) |
| Other | 0.576 | 0.106 | 0.318 | 0.09 (66) |
| Total | 0.551 | 0.154 | 0.294 | 1.00 (751) |

Table 2 illustrates how adjusting the confidence threshold affected diagnostic accuracy and adoption rates. At the default 60% threshold covering all cases, the rate of ‘Excellent’ predictions remained at 55.1%, while ‘Excellent + Good’ constituted 70.6% of cases. Crucially, the false positive rate (misdiagnosis) was 29.4%. As shown in Figure 2, increasing the confidence threshold led to improved precision, but a decrease in adoption rates. Setting a strict 90% threshold reduced the misdiagnosis rate to 14.1%, effectively halving diagnostic errors. Concurrently, the ‘Excellent’ rate rose to 72.9%, though this came at the cost of analyzing only 47% of cases meeting this high-confidence criteria.

**Table 2.** Diagnostic Accuracy of GPT-4 Turbo at Varying Confidence Thresholds. This table shows the GPT-4 Turbo’s diagnostic accuracy and adoption rate at various confidence levels.

| **Confidence Threshold** | **Excellent (%)** | **Good (%)** | **Insufficient (%)** | **Adoption Rate** |
| --- | --- | --- | --- | --- |
| **≥ 60%** | **55.1** | **15.5** | **29.4** | **100% (751/751)** |
| **≥ 70%** | **55.3** | **15.5** | **29.2** | **99% (746/751)** |
| **≥ 80%** | **57.9** | **15.8** | **26.3** | **92% (689/751)** |
| **≥ 90%** | **72.9** | **13.0** | **14.1** | **47% (354/751)** |
| **= 100%** | **87.5** | **12.5** | **0.0** | **1% (8/751)** |

**Figure 2.**
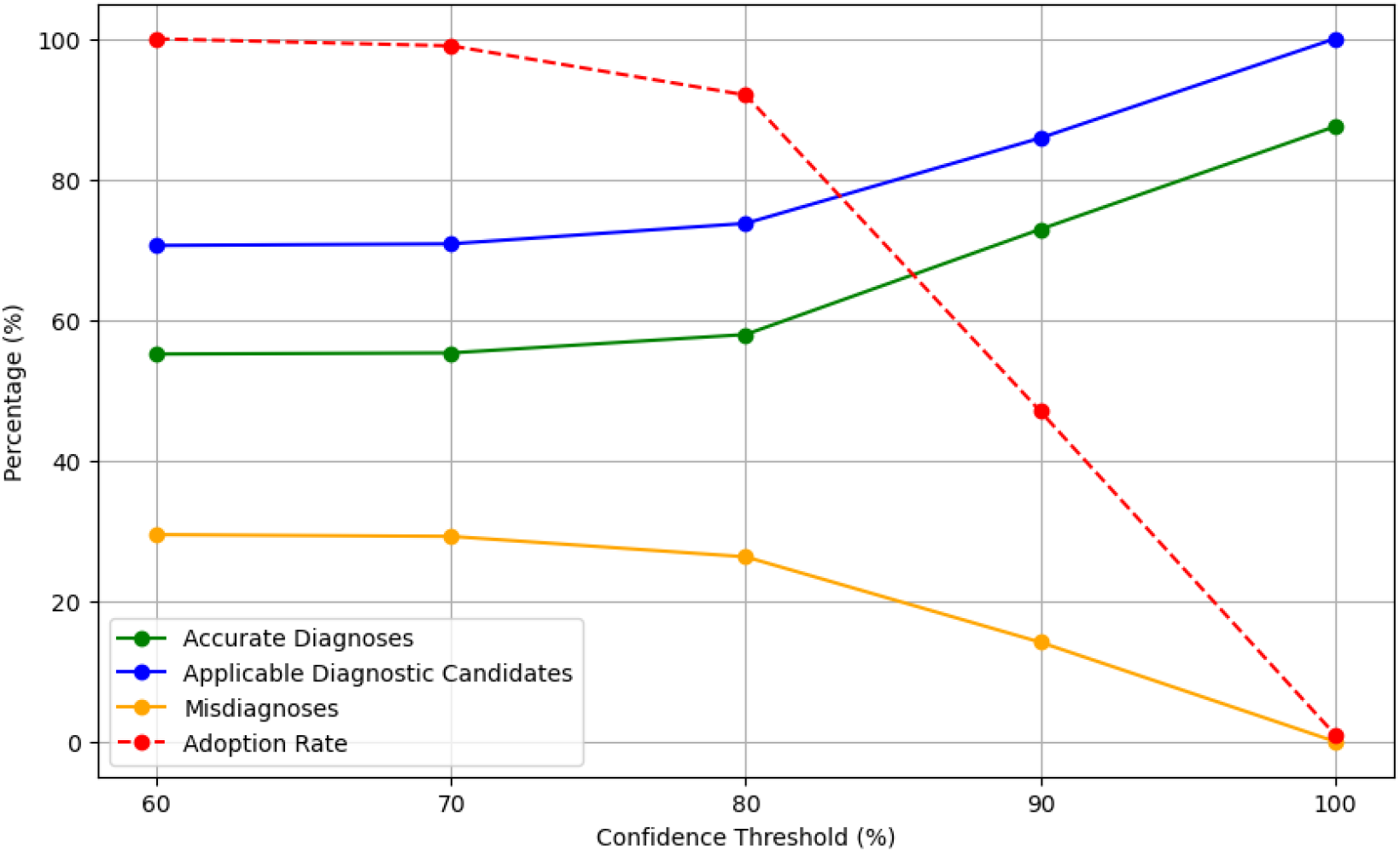
Impact of Confidence Thresholds on GPT-4 Turbo Diagnostic Performance This graph shows GPT-4 Turbo’s diagnostic accuracy across confidence thresholds. The green line shows accurate top suggestions that match the true diagnosis. The blue line indicates the cases with the correct diagnosis among the top five suggestions, suggesting usefulness. The orange line represents misdiagnoses, decreasing with higher confidence thresholds. The red dotted line shows the adoption rate, indicating how often the suggestions were suitable for clinical use. As confidence increases, so does accuracy, but adoption rates decline, highlighting a precision-practicality trade-off.

In summary, prompt engineering strategies coupled with confidence thresholds demonstrated a trade-off between diagnostic precision and practical utility for GPT-4 Turbo in neuroradiology cases. Higher confidence filters minimized misdiagnosis risks, but limited the proportion of cases the AI system could confidently assess.

## Discussion

### Improvement in Diagnostic Accuracy Through Prompt Engineering

The baseline performance of GPT-4 Turbo, marked with a 55.1% accuracy rate, is consistent with the results of previous studies in this domain, underscoring the ongoing challenge of improving diagnostic precision in complex medical fields (12). A strategic innovation in this study involved the incorporation of regulation in the prompt design to increase the utility of LLM responses. By shifting from presenting a single diagnostic candidate to offering a set of five, researchers increased the likelihood of including the correct diagnosis within LLM suggestions to 70%, consequently reducing the rate of insufficient responses to 30%.

### Risk Reduction through Confidence Threshold

We introduced a novel approach to using confidence thresholds to filter LLM responses, adding an analytical layer. By establishing a high confidence threshold (90% or higher), the probability that LLM provided an inaccurate diagnosis was effectively halved from 29.4% to 14.1%. This highlights the potential of prompt engineering to increase the utility of LLM in clinical settings. The quantitative impact of employing confidence thresholds is substantial, and the number of insufficient responses from LLMs in a dataset of 751 cases is expected to decrease from 221 to 50. However, this strategy reveals a significant trade-off that requires human decision-making in the absence of LLM suggestions in 53% of cases.

### Consideration and Limitations

The efficacy of diagnostic efforts is fundamentally based on human experience. The performance of LLMs in diagnostic tasks is contingent upon several factors: the quality and quantity of clinical information and imaging findings provided, the capability to extract pertinent insights from images, and selecting the most probable diagnosis from the LLM’s suggestions. Although setting high confidence thresholds significantly mitigates the risk of incorrect diagnoses, it concurrently limits the LLM’s applicability by reducing the number of cases it can address. Additionally, asking medical professionals to consider multiple diagnostic proposals could potentially improve the precision of the diagnosis. However, this approach introduces an additional cognitive load, which can affect clinical workflows and decision-making processes. A limitation of this study is its focus on evaluating precision in scenarios where sufficient information is provided for diagnosis. There is a need for further research on the LLM’s performance with real-world data, which may present incomplete information or atypical cases, and in domains beyond neuroradiology.

### Future Perspectives

The results of this study suggest avenues for enhancing the accuracy of LLM suggestions:

* Expanding the learning data, especially for tasks where confidence levels were low.
* Implementing few-shot learning as part of prompt engineering to refine the LLM reasoning process by providing it with examples of human thought processes.

Discussing the potential to improve these methods to increase the proportion of high-confidence cases is crucial. Future studies could explore optimizing these strategies to balance diagnostic precision with usability, possibly integrating LLM suggestions more effectively into clinical decision support systems.

## Conclusion

Prompt engineering strategies and confidence level thresholds applied to GPT-4 Turbo improve the diagnostic accuracy in neuroradiology, offering a promising avenue for AI-assisted clinical decision-making.

## Data Availability

All data produced in the present study are available upon reasonable request to the authors

## ACKNOWLEDGMENTS

This work was supported by the Japan Society for the Promotion of Science (JSPS) KAKENHI under Grant [22K07674]. The authors would like to express their gratitude for the financial support provided, which has been instrumental in the advancement of this research.

## Notes

### Competing Interest Statement

The authors have declared no competing interest.

